# Exposure to Wood dust and its Respiratory Health Effects Among Wood Workers in Yaoundé (Cameroon)

**DOI:** 10.1101/2023.12.28.23300613

**Authors:** Innocent Takougang, Gilles Wilfried Bukam Guemnyen, Michel Franck Edzamba, Fabrice Zobel Lekeumo Cheuyem, Peguy Brice Assomo Ndemba, Walter Yone Pefura

**Affiliations:** Department of Public Health, Faculty of Medicine & Biomedical Sciences, The University of Yaoundé I, Yaoundé, Cameroon; Department of Physiology, Faculty of Medicine and Biomedical Sciences, The University of Yaoundé I, Yaoundé, Cameroon; Department of Internal Medicine and Specialties, Faculty of Medicine and Biomedical Sciences, The University of Yaoundé I, Yaoundé, Cameroon

**Keywords:** wood workers, exposure, respiratory symptoms, Cameroon

## Abstract

**Background:** Occupational respiratory diseases are responsible of one-third of all documented work-related deaths. Exposure to wood dust leads to multiple respiratory manifestations including cough, chest pain, asthma and altered lung function. The aim of this study was to assess the level of exposure to wood dust and its respiratory health correlates among woodworkers in Yaoundé.

**Methodology:** The present descriptive cross-sectional study was conducted in 37 carpentry workshops in the city of Yaoundé. From October 2021 to February 2022. The geographical site selection was purposeful, taking into account areas with large numbers of workers and woodworker shop. Woodworker shops were enumerated and randomly selected. Respiratory manifestations were assessed upon a clinical respiratory examination involving cough, expectoration, wheezing and dyspnea. The force expiratory volume was determined using a dry spirometer. Statistical analyses were performed using IBM SPSS version 23.0 software; tables and graphs were generated using Excel 2013 software. The significance level was set at 0.05.

**Results:** The study population was exclusively male, with a mean age of 34.04 ± 11.69 years. With 15.02 ± 12 years of woodwork experience. The respiratory symptoms reported were cough that was productive (41.8%) or dry (33.6%), chest pain (34.4%), dyspnea (41%) and wheezing (15.6%). The lung function decreased among the duration of woodwork experience.

**Conclusion:** Respiratory manifestation among woodworkers were reported. And there is a urgent need to implement woodwork safety measures including education on exposure and adherence to protective measure.

## Introduction

Occupational respiratory diseases account for 30% of all registered work-related diseases [1]. About 300 million people are affected each year with 2.3 million related deaths [2]. Respiratory diseases represent 17.1% of work-related fatalities [3]. In addition, at least 2 million people are exposed to wood dust around the world every day [4–6].

Wood dust is released into the air during machine maintenance, cleaning and at all stages of woodworking. The amount of dust emitted depends on the type of material, the specific operation, the machine characteristics and the working environment [7]. Inhaled wood dusts range from 6% to 75% of the total wood aerosol [8,9]. Wood dust particles enter the human body mainly through the respiratory tract, causing direct toxicity by damaging alveolar, tracheal and bronchial epithelial cells. This leads to inflammation and irritation of the respiratory system, resulting in coughing, wheezing, chronic bronchitis, tightness of the chest, and asthma, alveolitis, and decline in lung function [4,10–12].

Studies suggest that workers in the wood-processing industry who are exposed to wood dust face a higher likelihood of developing respiratory symptoms [13–15]. Furthermore, the respiratory symptoms predominantly reported in association with wood dust exposure were wheezing, chest tightness, coughing, phlegm production, and dyspnea [16].

Research has demonstrated a positive correlation between lung function impairment and the extent of exposure to wood dust. Exposure to wood dust in the workplace results in decreased Forced Vital Capacity (FVC) and Forced Expiratory Volume (FEV) [6,14,17,18].

Guidelines and regulations regarding the exposure to wood dust are not frequently implemented in Low and Middle-Income Countries [6,19,20]. Additionally, there is a lack of extensive morbidity and mortality records related to occupational exposure to wood dust [19,21,22]. In Cameroon, woodworking provides the second highest number of jobs in the public sector (170,000) and is the country’s second largest export product after oil (10%). However, there is a lack of data regarding respiratory disorders caused by woodworking [23,24]. The present investigation aimed to assess the lung function impairments among woodworkers with varied levels of exposure to wood dust in Yaoundé.

## Methodology

The present cross-sectional descriptive woodwork study was carried out in two Yaoundé health districts from October 2021 to February 2022.

Workers with a history of recent abdominal or chest surgery, heart failure, TB, emphysema, and acute illness were all excluded from the lung function test.

The selection of study participants was based on a multi-stage sampling technique. Two of the eight health districts of the city of Yaoundé were purposefully selected for hosting the main wood processing establishments. All woodworkers who consented to participate were included.

Wood factories characteristics were found using some items like size, type of machine used, dust control and dust removal method. The level and duration of exposure to wood dust was assessed using an adapted version of the American Thoracic Society questionnaire (ATS) [24]. The questionnaire items related to socio-professional characteristics (age, gender, work experience). A clinical respiratory examination as medical history (tuberculosis, asthma, pneumonia, smoking) was then conducted and respiratory symptoms (chest pain, cough, dyspnea, wheezing) assessed. All interviews and clinical examination were carried out in secured private location by the work place.

Lung function assessment was performed using a dry spirometer (Vitalograph1, Buckingham, UK) [25]. Spirometry examinations were performed early in the morning, before the workers would start work. The technique was demonstrated to each participant prior to testing under supervision. FVC and FEV1 were assessed for each participant. Participant’s weight (kg) was measured using a standardized electronic weighing machine while they stood and wore light clothing. Height (cm) was measured using a portable stadiometer.

Statistical analyses were performed using IBM SPSS version 23.0 software. The FEV1/FVC were computed and used as an indicator of respiratory capacity. A simple linear regression was used to visualize lung function progression with year of exposure. A two-sided *p*-value˂0.05 was considered statistically significant.

The present investigations received ethical approval of the institution Ethical Review Committee of the Faculty of Medicine and Biomedical Sciences, the Regional Ethical Review Committee of Center Region. All examinations were under the supervision of a renowned pneumologist who ensured that all practices respected technical standards.

## Results

The present investigations involved 122 woodworkers from 37 selected factories.

More than half of wood working sites were of medium size (59.5%). The commonly used machines included sawing (86.5%), planing (94.6%). Almost two-third of wood factories used dry sweeping to remove wood dust (64.9%) (Table 1).

**Tableau 1.** Characteristics of studied wood factories in Yaoundé, 2021 (*n*=37)

| Variable | Count(n) | Frequency (%) |
| --- | --- | --- |
| **Wood factory size** |  |  |
| Small | 11 | 29.7 |
| Medium | 22 | 59.5 |
| Large | 4 | 10.8 |
| **Type of machine used** |  |  |
| Sawing machine | 32 | 86.5 |
| Planing | 35 | 94.6 |
| Sanding | 28 | 75.7 |
| Band saw | 10 | 30.3 |
| Degreaser | 5 | 15.2 |
| Portable electric tools | 3 | 8.8 |
| Mortising machine | 2 | 5.9 |
| Drilling machine | 4 | 12.1 |
| **Dust removal method** |  |  |
| Dry sweeping | 24 | 64.9 |
| Compressed air | 13 | 35.1 |
| **Dust control** |  |  |
| Yes | 2 | 5.4 |
| No | 35 | 94.6 |

### Socio-professional Characteristics of woodworkers

All study participants were male aged 30-39 years (48.4%) with less of than 10 years of professional experience. Most of woodworkers were indiscriminately involved in sanding (96.7%), wake (86.9%) and planing (84.4%) (Table 2).

**Tableau 2.** Socio-professional characteristics of Yaoundé wood factory employees, 2021 (*n=*122)

| Variable | Count (n) | Frequency (%) |
| --- | --- | --- |
| **Age (years)** |  |  |
| Under 20 | 9 | 7.3 |
| 20-29 | 44 | 36.0 |
| 30-49 | 59 | 48.3 |
| Over 50 | 10 | 8.1 |
| **Working post** |  |  |
| Wake | 106 | 86.9 |
| Planing | 103 | 84.4 |
| Sanding | 118 | 96.7 |
| Varnishing | 96 | 78.7 |
| **Professional experience** |  |  |
| Less than 10 years | 53 | 43.4 |
| 10-20 years | 24 | 19.6 |
| More than 20 years | 45 | 36.8 |

### Respiratory Symptoms

The most reported respiratory symptoms were chronic cough (75.4%), dyspnea (40.9%) and chest pain (34.4%) (Figure 1).

**Figure 1:**
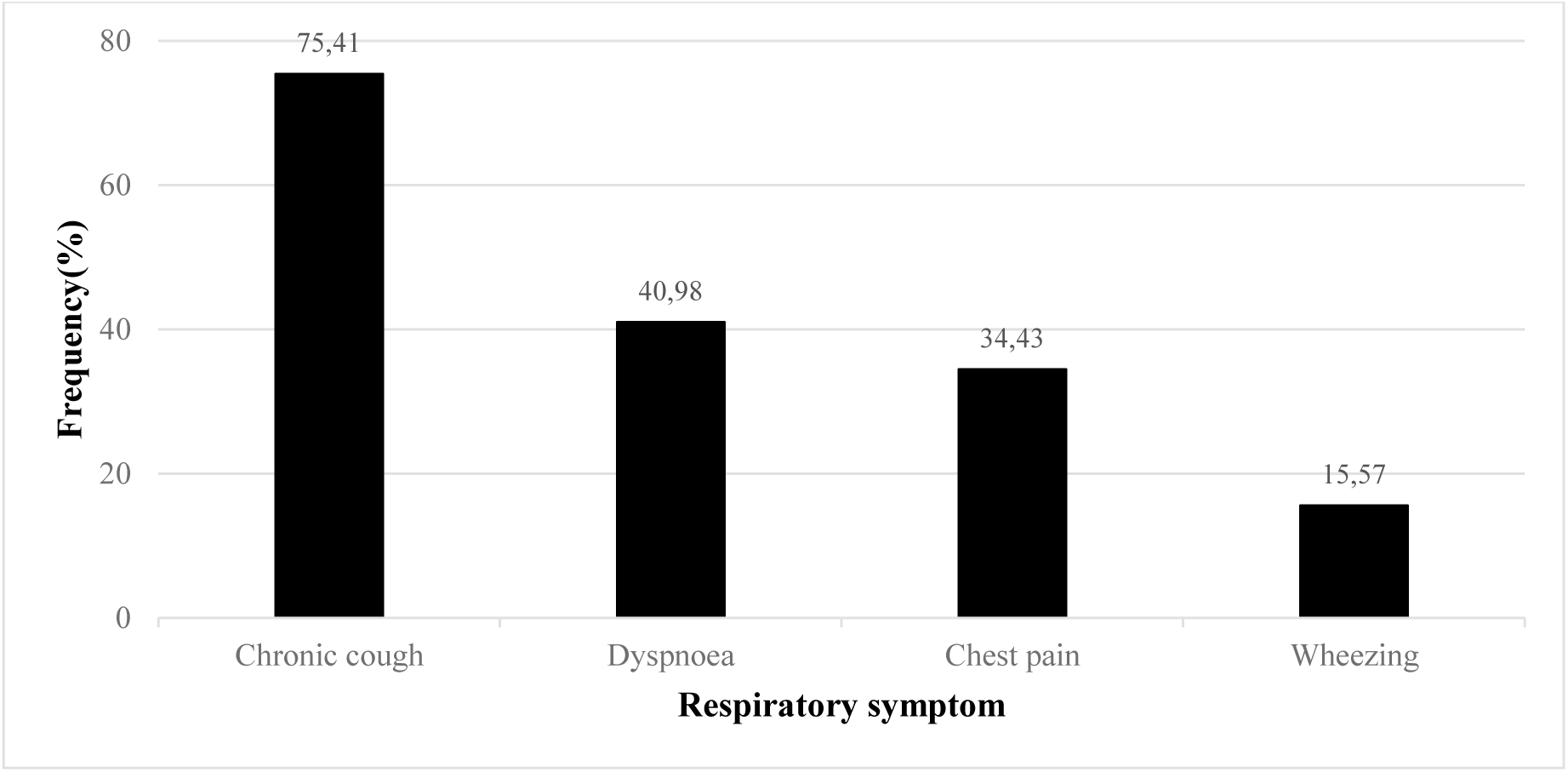
Respiratory symptoms among wood workers in Yaoundé, 2021 (*n=*122)

Unit separation did not exist for more than two-third of wood factories (73%) and all of them applied no restriction of pollution areas.

Participants reported a past history of pneumonia (23.7%), smoking (22.1%) and tuberculosis (3%) (Table 3).

**Tableau 3.** Reported history of respiratory diseases among wood worker in Yaoundé, 2021 (*n=*122)

| Past history | Count (n) | Frequency (%) |
| --- | --- | --- |
| Tuberculosis | 4 | 3 |
| Asthma | 10 | 8.2 |
| Pneumonia | 29 | 23.7 |
| Current smoker | 6 | 4.9 |
| Ex-smoker | 21 | 17.2 |

Almost one-third of workers declared using of personal protective devices (PPD) against wood dust (30.3%). Meanwhile more than half (PPD) user reported a weekly recycling frequency (57.4%) (Table 4.)

**Table 4.** Personal protective equipment use and recycling frequency among wood workers in Yaoundé, 2021.

| Variable | Count (n) | Frequency (%) |
| --- | --- | --- |
| **Protective equipment** |  |  |
| Simple mask | 27 | 22.1 |
| Hood | 10 | 8.2 |
| None | 85 | 69.7 |
| Total | 122 | 100 |
| **Recycling frequency (Days)** |  |  |
| Never | 2 | 5.4 |
| 1 | 8 | 21.6 |
| 2-3 | 5 | 13.5 |
| 4-7 | 12 | 32.4 |
| More than 7 | 10 | 27 |
| Total | 37 | 100 |

### Spirometry

The forced vital capacity (FCV) declined with duration of exposure. The FCV decreases at a rate of 0.03 liters per year among Yaoundé wood workers (*p-*value= 0.03) (Figure 2).

**Figure 2.**
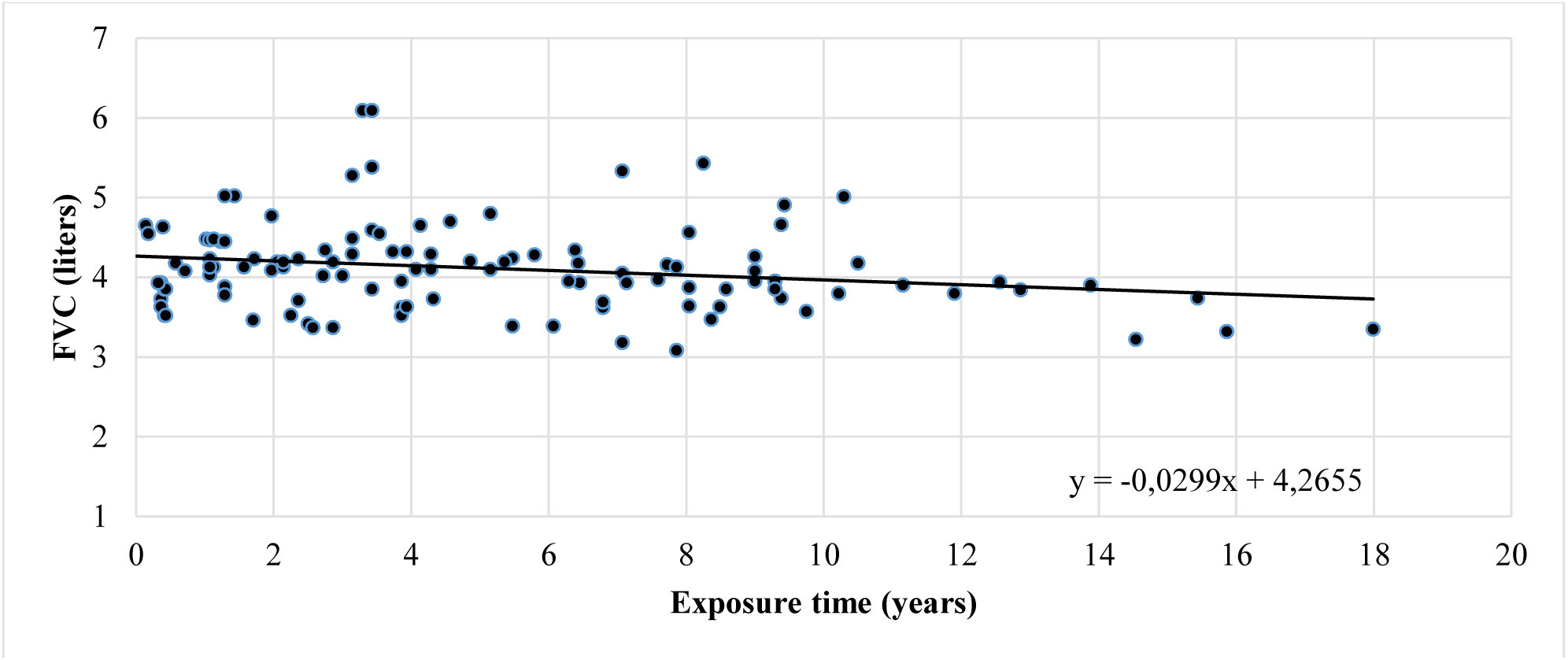
forced vital capacity (FVC) by duration of exposure to wood dust among woodworkers in Yaoundé, 2021, (*n=*122)

The FEV1 value decreases gradually with duration of exposition. A drop of 0.04 liters per year among wood workers (*p-*value= 0.02) (Figure 3).

**Figure 3:**
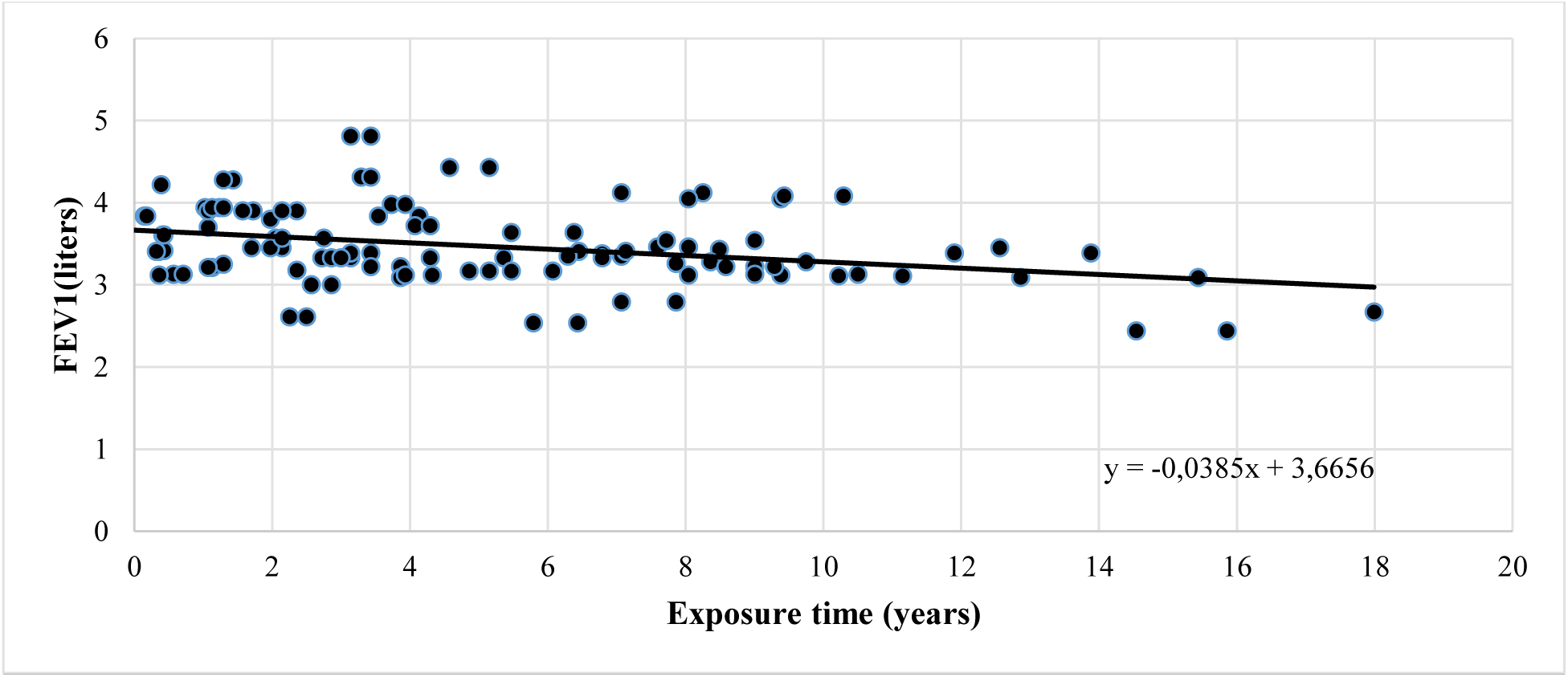
F*orced expiratory* v*olume in one second (FEV1)* by duration of *exposure* among wood workers, 221, (n=122)

The Tiffeneau ratio value decreased at an annual rate of 0.35 % among wood workers (*p-*value= 0.045) (Figure 4).

**Figure 4.**
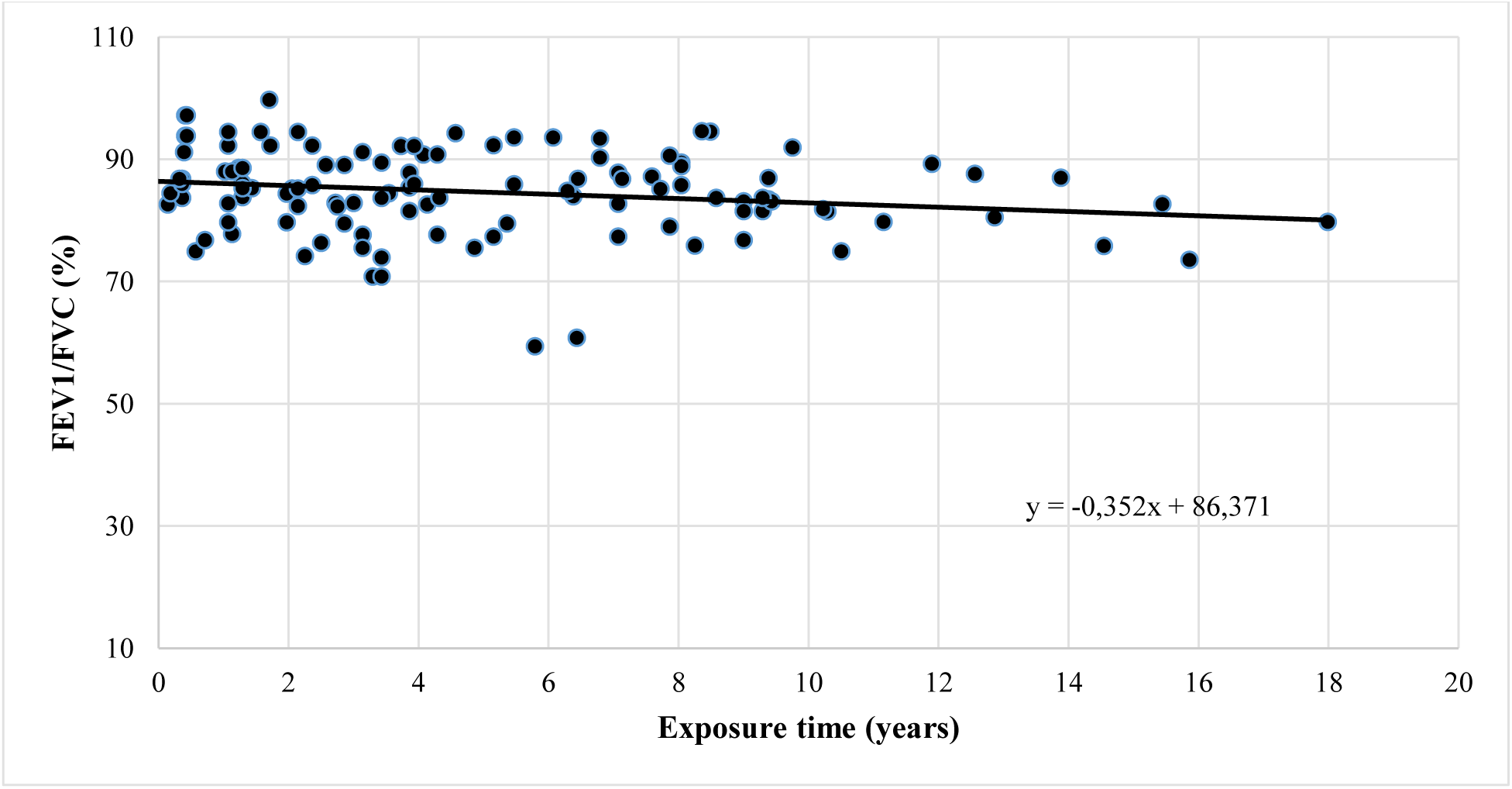
Tiffeneau ratio value and duration of exposure among wood worker in Yaoundé, 2021 (n=122)

## Discussion

Exposure to wood dust among woodworkers lead to a higher prevalence of respiratory problems. This could be due to poor adherence to guidelines and regulations related to occupation health and safety measures including the systematic use of PPD in the workplace. The concentration of wood dust and the duration of exposure influence the rate of occurrence of respiratory disorders among exposed workers. Specifically, the study found a high prevalence of chronic cough (75.4%), dyspnea (40.9%) and chest pain (34.4%). This might be due to the high concentration of wood dust found in the working area causing a chronic inflammation from continuous exposure to the wood dust. This result corroborates observation in Nigeria and Ethiopia [20,26]. A reduced occurrence of respiratory symptoms was found in woodworkers who worked in well-equipped environments and adhered to the enforced use of personal protective equipment (PPE), such as masks [27]. Additionally, these workers manifested a high level of knowledge and a positive attitude towards dust prevention measures.

### Contributing factors

The use of respiratory protective equipment is a key factor in occupational health and safety for woodworkers. These study findings outlined a poor observance of personal protective measures because, more than half of workers did not use any PPE as recommended. Such behavior are described where no regulation regarding the wood processing activities are implemented [19]. There was a clear absence of a separation between areas that are polluting and those that are not. Furthermore, it was observed that the factories did not have any restrictions on polluting areas for essential workers and there were no enclosures for machinery that produces wood dust. Additionally, no vacuum system was employed for dust elimination. Because poor working conditions could increase the risk of respiratory impairment, as the connection between inadequate adherence to recommend wood working regulations and increased dust exposure among workers has been well established in other studies [9]. Previous respiratory diseases such as those identified may contribute to the onset of current respiratory symptoms. Other studies have reported that workers who had a previous respiratory disease were 3.9 times more likely to experience respiratory symptoms [20]. Additionally, numerous studies have reported a significant association between work duration and the presence of at least one respiratory symptom. This could be due to the fact that extended exposure in the workplace leads to the highest levels of dust accumulation in the respiratory system [20,26,28–30].

### Respiratory performance

The average respiratory parameters in this study were lower than normal expected values. This indicates that woodworkers may experience reduced respiratory function due to the high concentration of inhaled particles (mainly wood dust), reflected in the number of workers with pulmonary function impairment. Woodworking activities combined with a lack of protective equipment increases exposure to respirable wood dust. This finding suggests that exposure to wood dust in the workplace may lead to a decline in lower Forced Vital Capacity (FVC) and Forced Expiratory Volume in one second (FEV1) and ratio of FEV1/FVC, consistent with prior research carried out in other regions of Africa [6,19]. As the duration of exposure to wood dust increased, the lung function index decreased resulting in increased respiratory disorders. This result corroborates observation in in Ghana and Pakistan [19,31].

## Conclusion

Woodworkers may experience various respiratory symptoms such as coughing, chest tightness, shortness of breath and wheezing. Contributing factors include prior respiratory conditions, inappropriate use of personal protective equipment, restrictions on pollution zones and the length of exposure to wood dust. Lung function deteriorates with increased exposure to wood dust. It is necessary to introduce health surveillance for workers and enhance the work environment to mitigate the risk of developing respiratory ailments.

## Limitation

This was a descriptive cross-sectional study, so cause and effect relationships could not be clearly established. The study did not measure the amount of wood dust each worker was exposed to during working hours. Workers may have a recall bias for respiratory symptoms, as symptoms are not always easy to identify or remember. The results should therefore be considered as preliminary measures that provide a starting point for further research, including cohort studies generate stronger evidence of causative relationship between health events.

## Data Availability

All data produced in the present work are contained in the manuscript

## Declaration

## Author’s Contribution

## Ethical Approval Statement

The protocol was approved by Institutional Review Board (IRB) of the Faculty of Medicine and Biomedical Sciences of Yaoundé and the ethical clearance: N°147/UY1/FMS/VDRC/DAASR/CSD granted. Informed consent was obtained from participants prior to inclusion in the study.

## Consent for publication

All participants gave their informed consent for publication of final results.

## Availability of data and materials

All data generated or analyzed during this study are included in this published article.

## Authors’ Contribution

Drafting of the study protocol, data collection, analysis and interpretation: G.W.B.G; Drafting, editing of manuscript: M.F.E.; Critical revision of the manuscript: F.Z.L.C., P.B.A.N. and W.A.Y.; Conception, design and supervision of research protocol and implementation, data analysis plan, revision, editing and final validation of the manuscript: I.T.

## Acknowledgements

Our gratitude goes to wood workers who accepted to participated to this study.

